# Identification and Modeling of Necroptosis-Related Genes Associated with the Prognosis of Breast Cancer

**DOI:** 10.1101/2024.08.02.24311442

**Authors:** Yukun Wen

## Abstract

**Background:** As one of the major cancers threatening human health, Breast Cancer (BC) has become the health concern of WHO (World Health Organization) all the year round. In recent years, new cases of BC have gradually increased, reaching 11.7% in 2020. In terms of treatment, the cell death is a basic way to treat cancer, and necroptosis is found to be a programmed form of necrotic cell death, which is related to cancer progression, metastasis and immune monitoring. In this study, the influence and role of Necroptosis-Related Genes (NRGs) in BC were analyzed, and the subtypes, prognostic model and subgroups were studied, respectively.

**Methods:** Four aspects were included in the research content. 1) Difference analysis. The Wilcoxon Test was applied to identify differences between normal people and BC patients. 2) Sub-type analysis. Based on Cox regression analysis, the key genes related to prognosis were extracted and applied to the Consensus clustering technology. Subsequently, after obtaining the subtypes, the Wilcoxon Test was applied to extract the differential genes of subtypes. 3) Prognostic analysis. Further, according to the survival time and state of patients, the genes related to the severity of the disease were extracted by the Cox regression, and the classification modeling of high- and low-risk was carried out by Lasso. 4) Sub-group analysis. Combined with the high- and low-risk labels of patients, the composition of differential genes was further analyzed. Subsequently, GO, KEGG, ssGSEA analyses were performed separately.

**Results:**

1) There are differences in gene expression between normal and BC patients. The results showed that, PLK1, CDKN2A and TERT were significantly different genes with |LogFC| > 2. In addition, PPI (Protein–Protein Interaction) demonstrated that CASP8, TRAF2, TNFRSF1A, HSP90AA1, CYLD, and FADD were hubs in the network. Moreover, co-expression relationship of these genes can be found in the correlation graph.
2) Unsupervised techniques suggested that there are 2 subtype characteristics in BC patients. The clustering results obtained the detailed clinical information of the 2 subtypes, and the survival analysis showed that different subtypes had different survival states. Similarly, the heat map also verified that these 2 types had different gene expression.
3) The validation demonstrated that the prognostic model has good effect. On the one hand, we found that ’BCL2’, ’FLT3’, and ’PLK1’ were the main genes with different expression levels in high- and low-risk patients. On the other hand, not only the ROC, risk curve and survival curve were verified, but also the PCA distribution and forest plots were demonstrated. These results showed that our model has good prognostic effect.
4) There were some differences in immune scores between high- and low-risk groups. A total of 94 genes were differentially expressed in different groups. Immune cell analysis and pathway analysis showed that, in general, immune scores of low-risk subgroup were higher than that in the high-risk subgroup.

**Conclusions:** Our findings revealed the crucial role of NRGs in BC. These are important for tumor immunity and can be used to predict the prognosis of BC.

## Introduction

Even today, BC is still a major public health problem and the second leading cause of cancer death in women^[1]^. Among women worldwide, the breast is the most prevalent of the five most common incident sites of cancer, accounting for 25.2% of the total, far higher than second, accounting for 9.2% of colon cancer^[2, 3]^. Not only women, the American Cancer Society estimated that in 2019 alone, there were about 2670 male patients, with a mortality rate of 18%^[4, 5]^. Therefore, the BC has become one of the most important cancers threatening global health.

On the other hand, cell death plays an important role in cancer treatment. Because most tumors have inherent apoptosis resistance, necroptosis has been gradually considered as a therapeutic strategy because of its mechanism of inducing other cell death^[6, 7]^. As a genetic programmed form of necrotic cell death and a form of regulated necrosis, necroptosis is a programmed lytic cell death pathway which provides new targets for therapeutic intervention^[8–10]^. In addition, apoptosis and necrosis are two main types of cell death with different cell morphology and pathways^[11]^. Although both are associated with inflammation, unlike apoptosis, necroptosis causes inflammatory responses by releasing damage-associated molecular patterns^[12]^. Furthermore, as a key cell killing mechanism, controlled by receptor interacting protein 1 (RIP1), RIP3, and mixed lineage kinase domain like (MLKL), necroptosis is related to cancer progression, metastasis and immune monitoring^[13–15]^. In general, more and more evidence reveals that necroptosis has a certain association with cancer treatment^[16–18]^.

In order to analyze the relationship between genes and diseases, this study takes BC disease as the research object, NRGs as the breakthrough point, and discusses the influence of related genes on BC in detail.

Four parts were included in this paper. Firstly, the different **NRGs** between normal people and patients was analyzed. Then, according to the screened genes related to prognosis, BC diseases were divided into 2 different subtypes. On this basis, the prognostic model was constructed and verified. Finally, the enrichment analysis and immunoassay were carried out based on different subgroups.

Overall, this study aims to identify the correlation between the expression of NRGs and the prognosis of BC patients, so as to establish a predictive model. We believe that this work has certain reference significance for the prognosis of BC and clinical guidance.

## Methods and Materials

### The Main Content and Process

The flowchart of this paper is shown in the Fig. 1. The process of this study can be divided into four main parts: Difference Analysis, Sub-type Analysis, Prognostic Analysis, and Sub-group Analysis. First, 67 NR BCGs (Necroptosis-Related Breast Cancer Genes) and 62 Different NRGs (Different Necroptosis-Related Genes) were extracted, and PPI, Correlation Network, Heatmap analysis were performed around them. Then, the Consensus clustering was conducted based on 12 Prognostic NRGs (Prognostic Necroptosis-Related Genes), patients were divided into 2 Subtypes, and differential analysis of clusters was performed.

**Figure 1.**
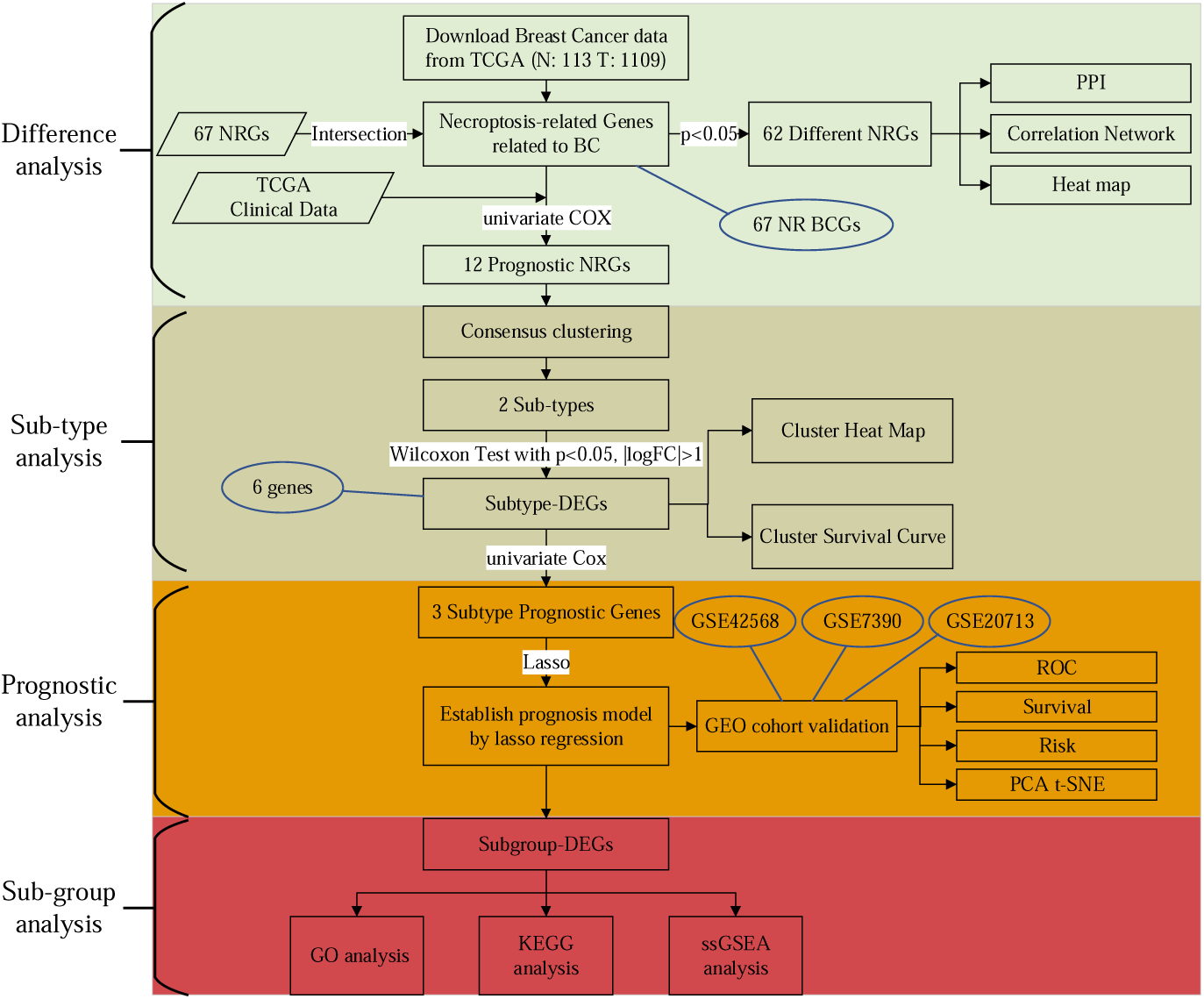
The work flow chart of this research.

Further, 6 **Subtype DEGs** (Subtype Different Expressed Genes), which were considered to be 2 subtypes with differences in patient survival, were screened. Next, the prognostic model was built through 3 **Subtype Prognostic Genes** by Lasso (Least Absolute Shrinkage and Selection Operator) Regression, and the model were tested in different aspects. Finally, **Subgroup DEGs** (Subgroup Different Expressed Genes), i.e., differential genes between different subgroups are extracted, and differential analysis were performed around high- and low-risk subgroups, such as GO, KEGG, and ssGSEA analysis.

### Collection of Data

The data of complete RNA sequencing (RNA-seq) and clinical characteristics of patients from 113 normal cases and 1109 BC patients were collected from the TCGA-BRCA project of TCGA (Texas Cotton Ginners’ Association) on 9 November 2021(https://portal.gdc.cancer.gov/repository). Data as validation sets are downloaded from GEO (Gene Expression Omnibus) databases (https://www.ncbi.nlm.nih.gov/geo/) with IDs of GSE42568, GSE7390, and GSE20713, respectively.

### Necroptosis-Related Genes

Previous studies have a certain basis for the NRGs. According to the published literature^[7]^, a total of 67 NRGs were obtained, of which 8 were obtained from the ‘necroptosis geneset M24779.gmt’ of GSEA (Gene Set Enrichment Analysis), and the other genes were collected from other literatures.

### Identification of Different NRGs, Subtype-DEGs and Subgroup-DEGs

A total of 62 Different NRGs were selected by the Wilcoxon Test, which set the threshold as *p* < 0.05. It should be noted that Different NRGs represent a set of genes with differences between normal people and patients. In addition, Wilcoxon Test method was also used in screening Subtype-DEGs and Subgroup-DEGs, in which the parameters were *p* < 0.05 and |LogFC| > 1.

### Protein-Protein Interaction (PPI) and Correlation Network analysis

In order to explore the relationship between Different NRGs at the protein level and to find Hub Genes, the PPI network was constructed with the Search Tool for the Retrieval of Interacting Genes (STRING), version 11.0 (https://string-db.org/). Besides, the minimum required interaction score for the PPI analysis was set at 0.9 (the highest confidence). In addition, the positive and negative correlations between genes were visualized by the Correlation Network analysis, indicating the interaction between genes.

### Univariate and Multivariate Cox Regression Analysis

The Cox regression, also known as proportional risk model, is a semi parametric regression model. The model takes the final state and survival time as the dependent variables, and can analyze the influence of many factors on survival time at the same time.

In this study, the Cox regression was applied to the screening of cancer genes, and it is employed as a key gene selection method for high- and low-risk. More specifically, by performing Cox analysis twice, the list of prognostic genes was obtained. For the first time, after obtaining the intersection NR BCGs of TCGA’s gene expression matrix and 67 NRGs, the Prognostic NRGs were obtained by the univariate Cox regression. For the second time, in the part of constructing prognostic model, Subtype Prognostic Genes were obtained by the Cox regression.

### Consensus Clustering

Clustering technology is an unsupervised machine learning method, which can divide objects into different categories. In this paper, patients were grouped into different subtypes by clustering to explore the differences of patients with different subtypes.

The Consensus clustering is a method to provide quantitative evidence for determining the number and members of possible clusters in the data set. It can not only divide the data into different subtypes, but also do not need to specify the number of categories artificially. Therefore, we performed Consensus clustering in the patient group with 12 Prognostic NRGs, and plotted survival curves for different subtypes of patients.

### Establishment and Validation of the Prognostic Model

The Lasso Regression is used to construct the prognostic model on Subtype Prognostic Genes. Lambda was used as the parameter of the method, and its interval was set to (− 1000, 1000). Through tenfold cross validation, it can be found that three genes were selected. Under these circumstances, the corresponding RSM was the smallest, and the model effect was the best. Therefore, these genes and their corresponding weights were selected to construct the model. The formula of the prognostic model is as follows:

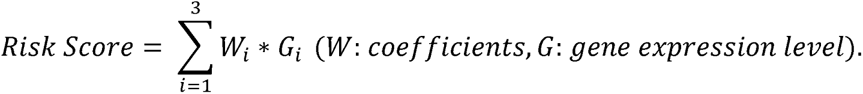

Each TCGA and GEO cohort patient’s corresponding Risk Score can be calculated by the prognostic model, and the patients can be divided into high- and low-risk subgroups by the median Risk Score in the TCGA cohort.

### The Survival, ROC, Risk, and PCA analysis

The R language (version, major: 4, minor: 1.1) was used to verify the prognostic model, including but not limited to survival analysis, ROC effect, risk analysis and PCA demonstration.

Specifically, 1) with the support of the software package “survival” and “surviviner”, the survival status of different subtypes was determined through the Kaplan Meier survival analysis. 2) In the correlation between the model prediction results and the real results, ROC is used for performance evaluation. Therefore, by introducing the “timeROC” package of R, the sensitivity and specificity of the prognostic model on BC overall survival at different years were evaluated by the ROC and AUC value. 3) The differences between subgroups were verified through risk analysis. 4) With the support of packages “Rtsne” and “ggplot2”, the distribution of different subgroups is effectively represented after PCA dimensionality reduction. In addition, the forest plots were drawn to simply and intuitively display single and aggregated research results through the "survival" package.

### Functional Enrichment Analysis of Subgroup-DEGs

By applying the Wilcoxon Test (|LogFC| > 1, p < 0.05) to TCGA cohort with high- and low-risk labels, **Subgroup-DEGs** can be obtained, which have significant differences on high- and low-risk subgroups. Then GO and KEGG analysis were applied to these Subgroup-DEGs to perform functional enrichment analysis.

### Immune Microenvironment Analysis

Immune microenvironment analysis can be used to analyze subgroup-DEGs in the immune microenvironment, and the ’GSVA’ of R package was used to obtain results related to immune cells and response.

### Statistical Analyses

All visualization and statistical analyses were performed using the R language (version, major: 4, minor: 1.1). The univariate and multivariate Cox regression was used to identify the independent prognostic factors. The Wilcoxon Test was used to extract the **Different NRGs**, **Subtype-DEGs**, and **Subgroup-DEGs**. The Kaplan–Meier method was used to compare the overall survival between low- and high-risk subgroups. The p < 0.05 was regarded to be significant.

## Results

### Identification of Different NRGs

This section identifies the Different NRGs in normal people and patients. The Wilcoxon Test method with *p* < 0.05 was set to find difference **NRGs**, and 62 **Different NRGs** were extracted from 67 **NR BCGs**. To explore the role of **Different NRGs**, the heatmap, PPI, and correlation analysis were carried out around these **Different NRGs**.

In the heatmap (As shown in Fig. 2 A), as the type label suggested, the blue means normal and the red means tumor, and different colors reflected different expressions of genes. Besides, ’***’ means *p* < 0.001, ’**’ means *p* < 0.01, and ’*’ means *p* < 0.05. Therefore, there were significant differences between normal and BC patients, and PLK1, CDKN2A, TERT were significantly different genes with |logFC| > 2.

**Figure 2.**
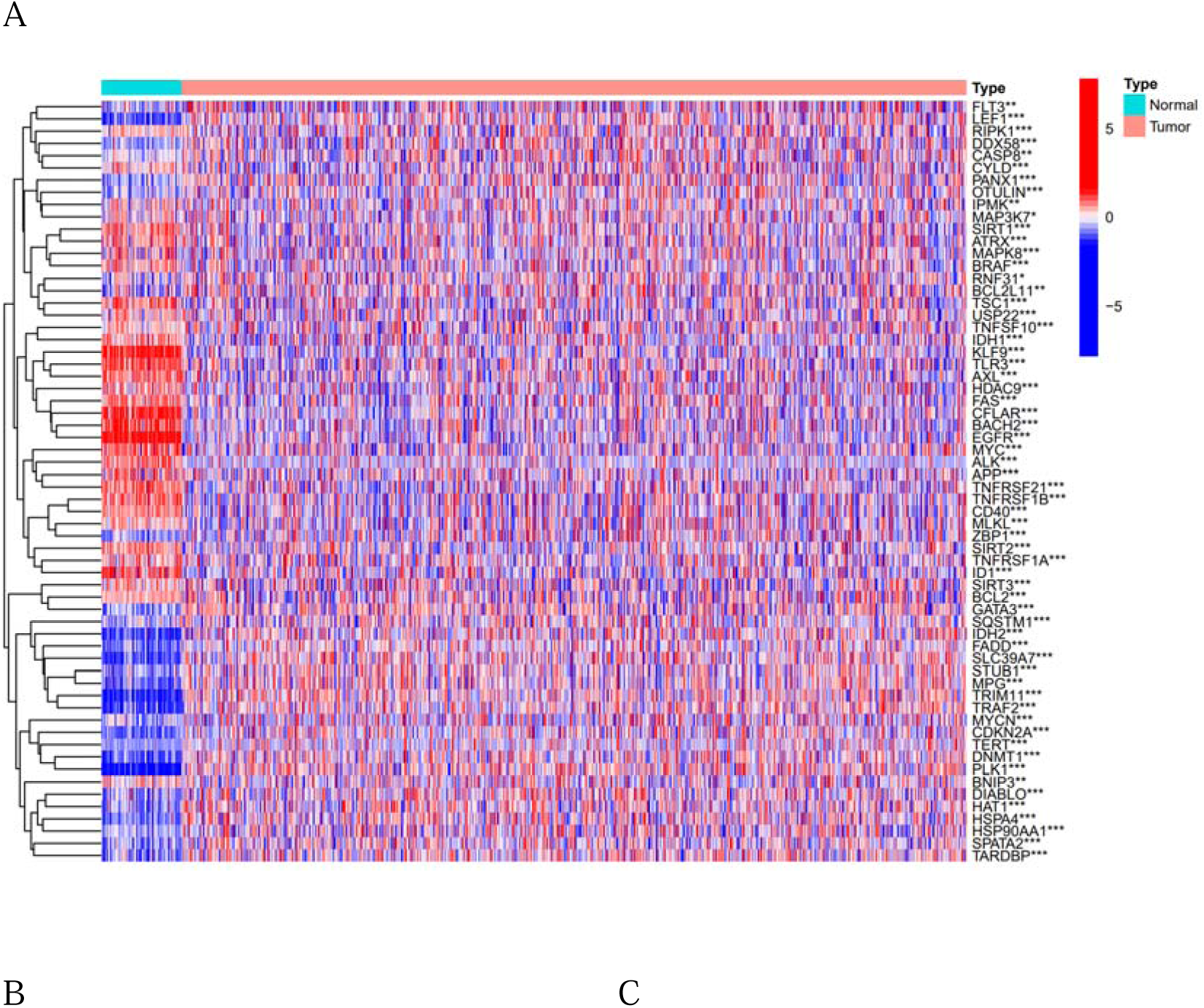
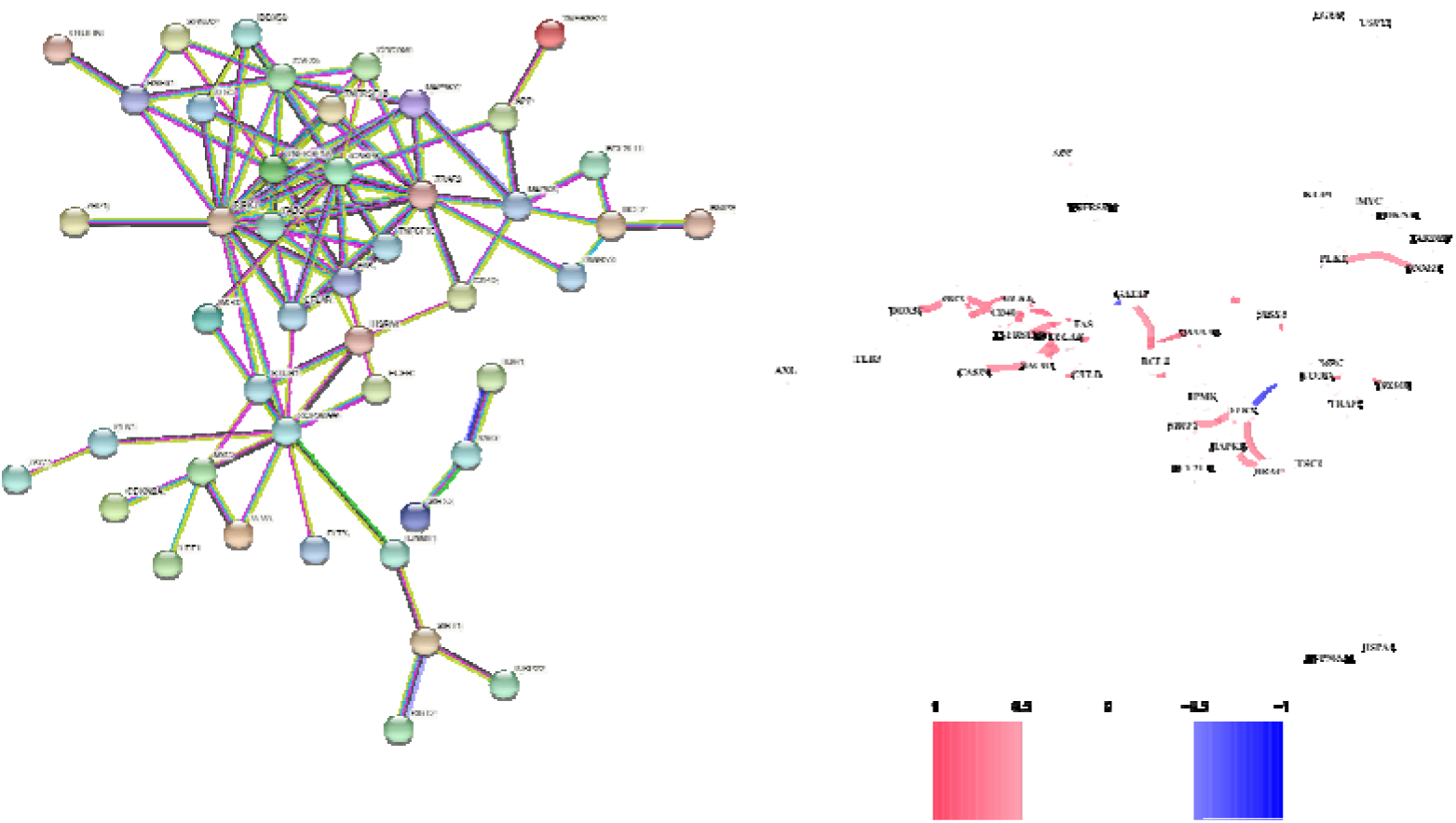
Expression and interaction of 62 Different NRGs. **A**: Heatmap of Different NRGs between normal tissue (N, blue) and tumor tissue (T, red) (blue: low expression level; red: high expression level). **B**: PPI network showed interaction of Different NRGs (interaction score = 0.9). **C**: The correlation network of Different NRGs (cutoff=0.35; red line: positive correlation; blue line: negative correlation. Color depth reflects correlation strength)

The PPI analysis (Fig. 2 B) showed that the RIPK1-centered proteins interact with each other under 0.900 confidence. In addition, CASP8, TRAF2, TNFRSF1A, HSP90AA1, CYLD, and FADD were also proteins with node degree > 8. Therefore, they were considered as hubs in PPI networks. Moreover, the correlation analysis (Fig. 2 C) showed the co-expression relationship among these genes, in which red and blue were positive and negative correlation respectively. For example, the MPG gene was positively correlated with STUB1 gene expression, while the MPG gene was negatively correlated with ATRX gene expression. The higher the correlation, the darker the color.

In addition, the univariate COX analysis was also applied to those 67 **NR BCGs**. In the cutoff of *p* < 0.05, 12 genes (’FASLG ’, ’IPMK’, ’TNFRSF1B’, ’PANX1’, ’BCL2’, ’FLT3’, ’PLK1’, ’BACH2’, ’HSP90AA1’, ’LEF1’, ’BNIP3’, ’CD40’) were found to be associated with prognosis, which were called **Prognostic NRGs**.

### Tumor Clustering based on DEGs

This section explores the results of patients being divided into different subtypes according to diagnostic NRGs.

By the Cox Regression, 12 **Prognostic NRGs** were obtained. In order to explore the influence of the above genes on BC subtypes, after deleting the normal people in the **gene expression matrix**, 1089 BC patients were clustered based on the **Prognostic NRGs**. By comparing the clustering results of different subtype numbers, 2 subtypes were finally decided (Fig. 3 A).

**Figure 3.**
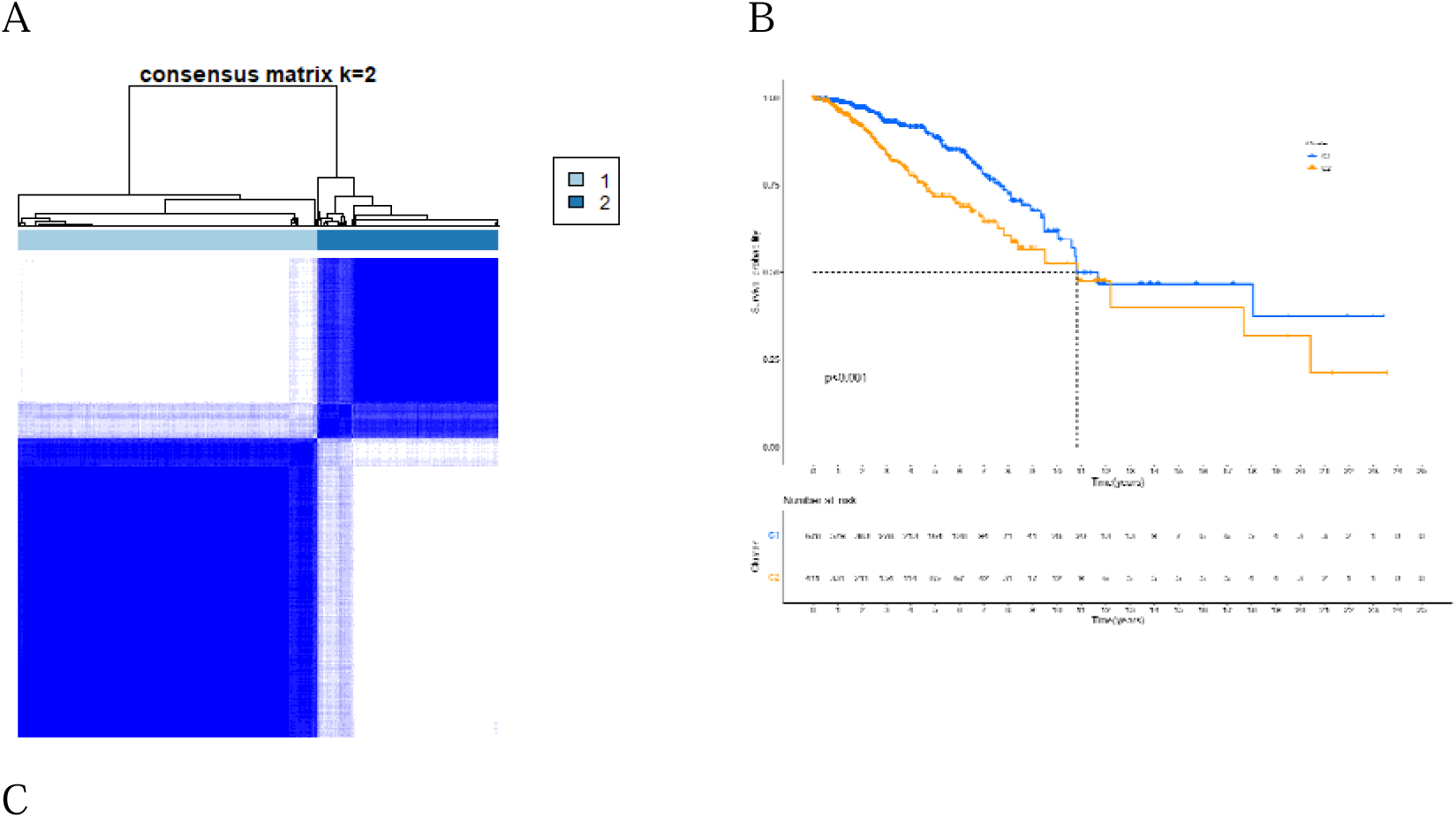
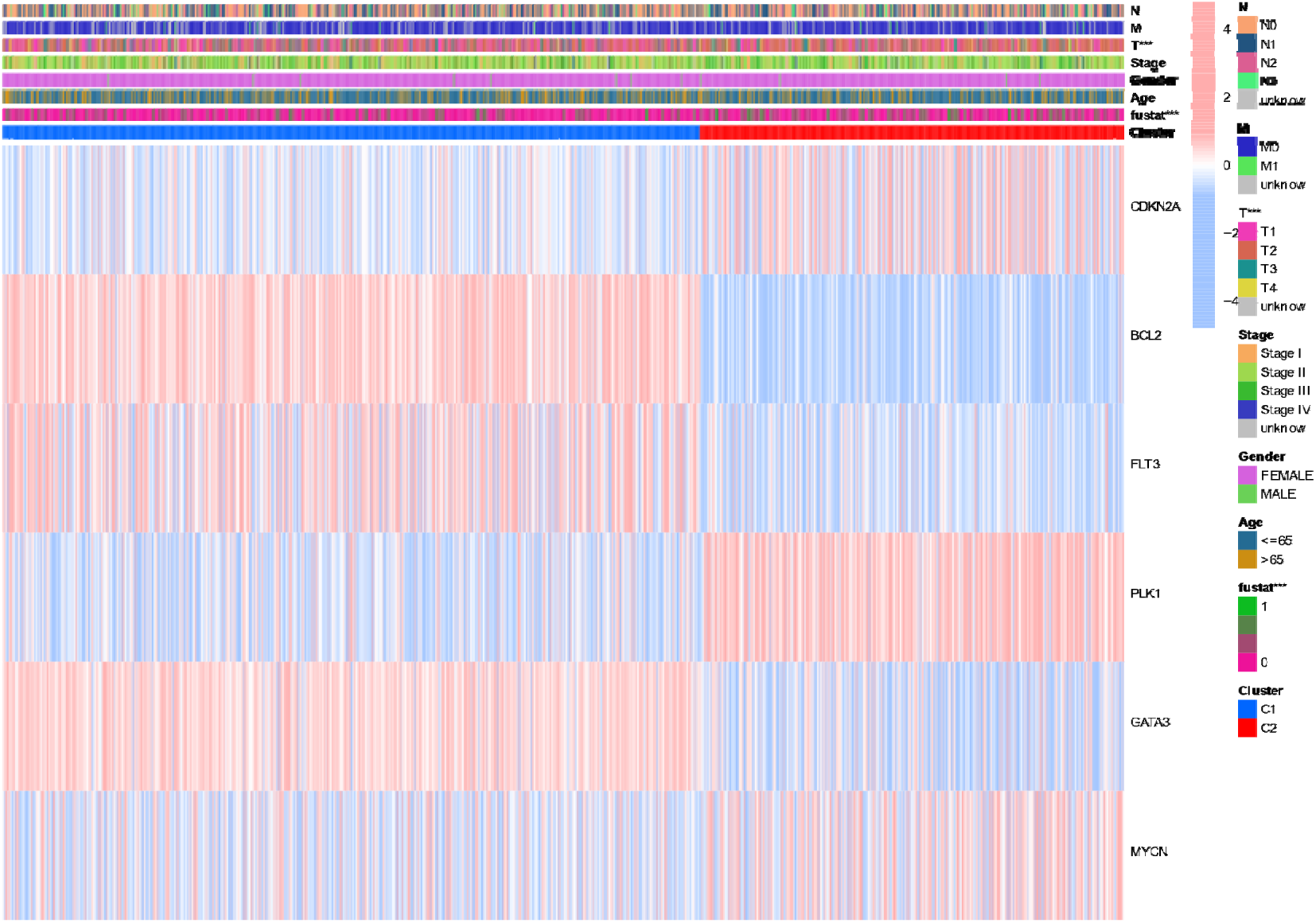
Tumor classification based on the NR BCGs. **A**: 1089 BC patients were grouped into 2 clusters according to the Consensus clustering matrix (minK = 2, maxK = 9). **B**: Overall survival curves for 2 subtypes by performing Kaplan–Meier method. **C**: Heatmap and the clinical features of 2 subtypes classified by the Subtype-DEGs (p value<0.05, “*”; p value<0.01, “**”; p value<0.001, “***”).

In addition, in order to explore the survival status between 2 different subtypes, the survival curve was drawn as shown in Fig. 3 B. The figure showed that patients with different subtypes have certain differences in survival status, it also reflects the reliability of the classification results.

After determining the clustering results, patients were assigned different subtype labels. Through the Wilcoxon Test, where the parameters were set to *p* < 0.05 and |LogFC| > 1, 6 genes were screened, which was called Subtype-DEGs and consisted by CDKN2A, BCL2, FLT3, PLK1, GATA3 and MYCN. The heat map was established by using the above genes. From the Fig. 3 C, we found that there were significant differences in the expression of these genes in different subtypes, and verified that they were the key genes to distinguish different subtypes.

### Establishment of Prognostic Gene Model

This section introduces the gene selection and modeling process of prognosis model. Based on the survival differences between 2 subtypes, the Wilcoxon Test was applied to 67 **NR BCGs**, and finally found that there were 6 genes with *p* < 0.05 and |LogFC| > 1, which indicated that they were related to prognosis.

After merging the data of TCGA and GEO corresponding to the 6 genes and removing the batch effect, we obtained the 6 **Same-EGs** and their corresponding expression levels. Then, these 6 **Same-EGs** were combined with the survival data of TCGA and GEO, respectively. Moreover, the univariate Cox analysis with *p* < 0.05 was performed on the combined TCGA **Same-EGs** and 3 **Subtype Prognostic Genes** were obtained, which were ’BCL2’, ’FLT3’, and ’PLK1’. Then, the Lasso Regression was applied to establish the prognostic model. The Lasso’s cvfit graph (Fig. 4A) showed that the model works best when the number of genes was 3. Besides, it can be seen in the Lasso’s lambda figure (Fig. 4B) that the convergence process of lambda and coefficients. Therefore, ‘BCL2’, ‘FLT3’, ‘PLK1’ and their corresponding weights were finally selected to build the prognostic model as follows:

**Figure 4.**
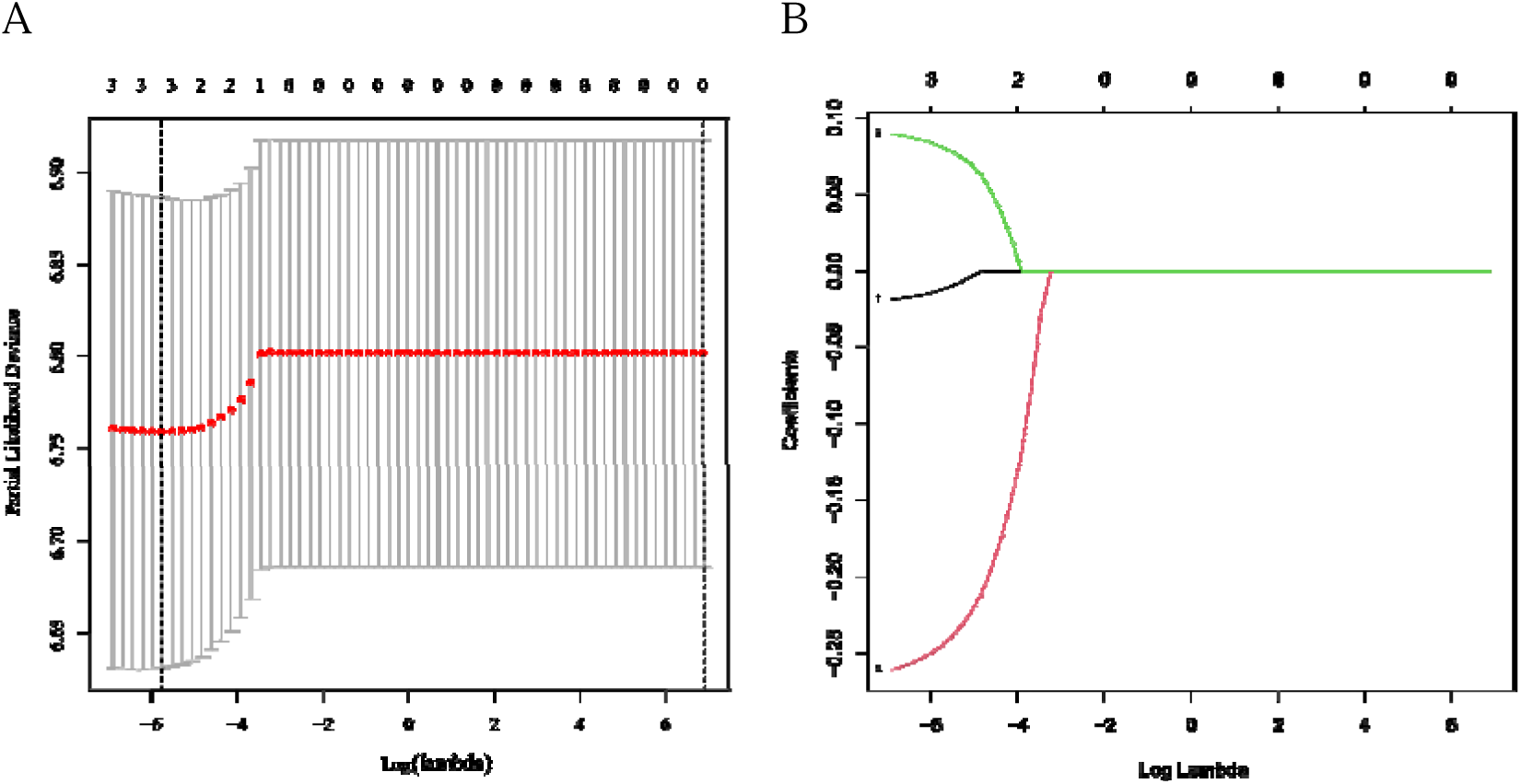
Lasso Regression model. **A**: The adjustment and selection of parameter lambda in lasso regression. **B**: Lasso regression of three key genes related to patient survival

The risk score for each patient in TCGA and GEO cohorts can be calculated according to the prognostic model. Since the risk score of the TCGA cohorts does not meet the normal distribution, the median (-0.09099686) was used as the dividing line of the high- and low-risk subgroup. Similarly, the medium value of the TCGA risk score was also applied to 3 GEO cohorts. In this way, TCGA and GEO patients can get the corresponding risk score and label.

### Evaluation and Analysis of the Prognostic Model

This section verifies the prognostic model in different aspects to confirm the prognostic effect.

In order to comprehensively analyze the model effect, the ROC effect, risk analysis, survival analysis, examination of PCA distribution, univariate and multivariate Cox analysis, and gene expression of different subgroups were discussed respectively. It should be noted that TCGA (T=1109, N=113) and GEO cohorts (GSE42568) were executed in all indicators, and GEO cohorts (GSE20713, GSE7390) were verified in the first four indicators.

ROC reflects the deviation between model prediction and reality, and is one of the indicators to reflect the accuracy of the prognostic model. Specifically, Fig. 5 (A1-B1) showed the ROC curves and AUC values of TCGA and GSE42568 cohorts in 1-, 3- and 5-years, while Fig. 5 (C1-D1) showed two other cohorts in 3-, 5- and 7-years.

**Figure 5.**
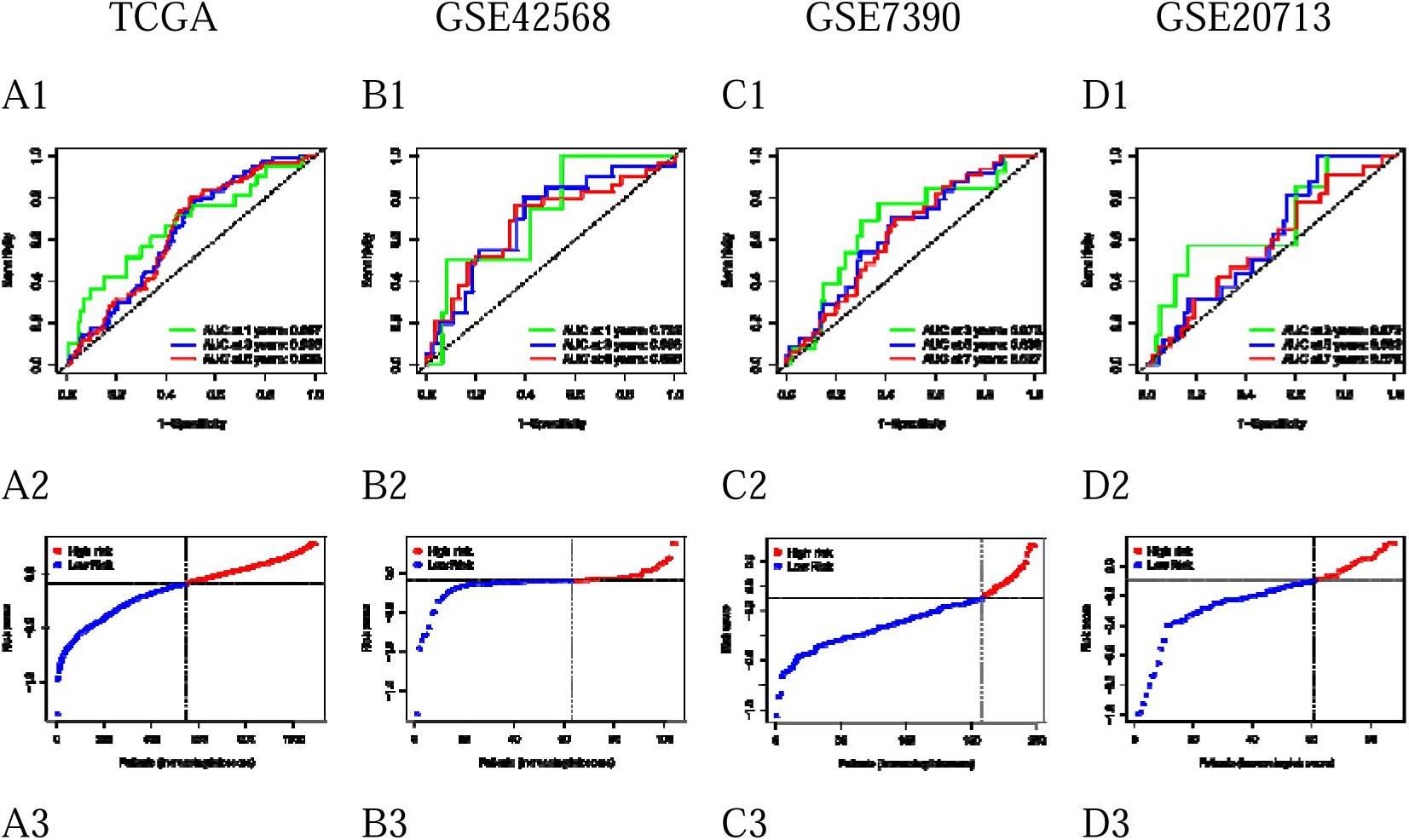
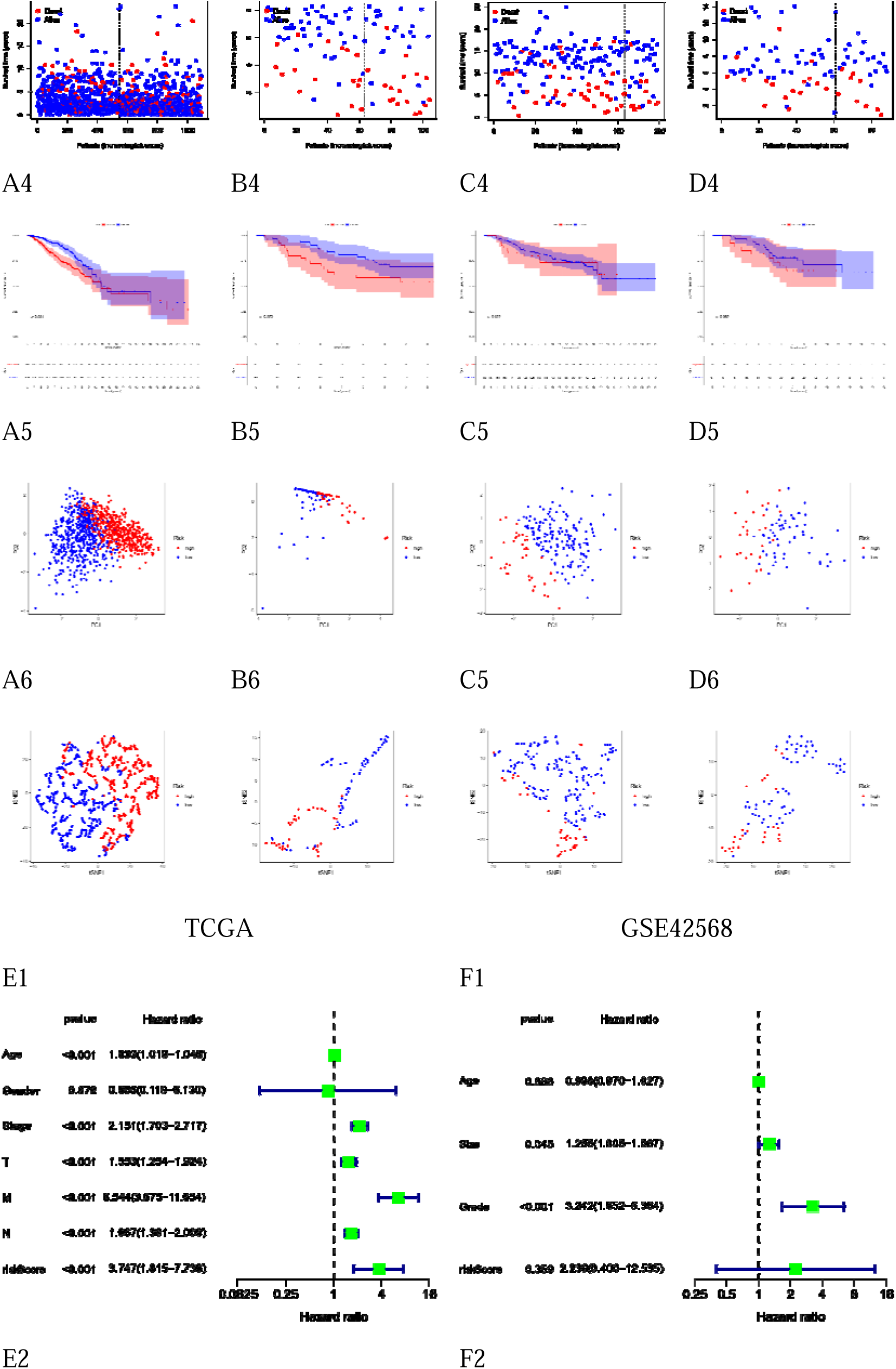
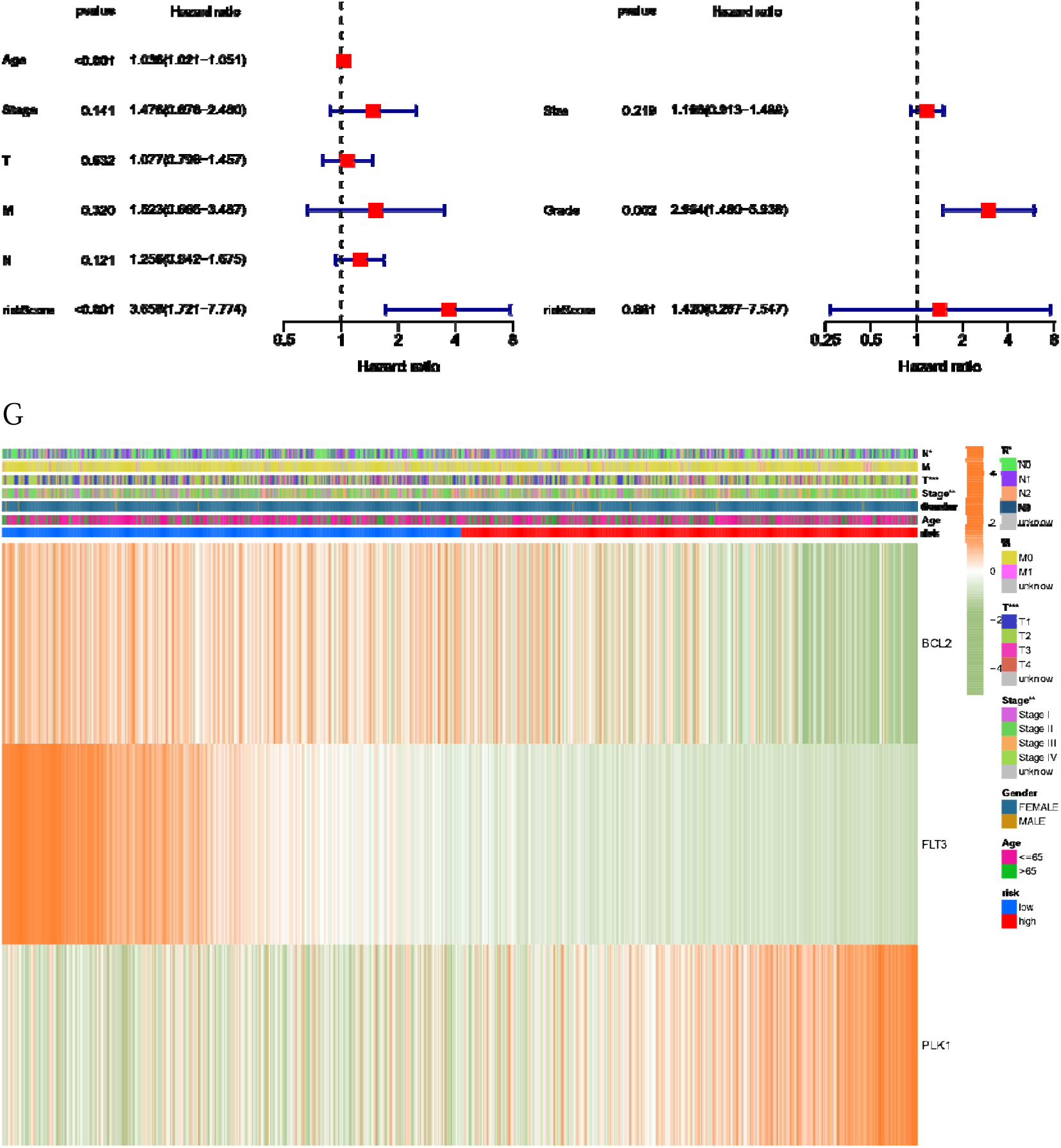
Validation of the prognostic model. **A**, **B**, **C**, and **D**: represent TCGA, GSE42568, GSE7390, and GSE20713 cohorts, respectively. **Numbers 1** to **6**: represent the corresponding ROC curve, risk curve, survival and death states graph, survival curve, PCA distribution graph, and t-SNE distribution graph, respectively. **E1** and **E2**: univariate Cox and multivariate Cox analysis of risk score for TCGA cohort, respectively. **F1** and **F2**: univariate Cox and multivariate Cox analysis of risk score for GSE42568 cohort, respectively. **G**: The heat map associated with the Subtype Prognostic Genes among subgroups.

The reason for choosing different years is that we can check the accuracy of AUC in different years. As can be seen A1-D1 from the Figure 5, the verification reflected that the model generally has high accuracy. For example, the AUC values of TCGA and GSE42568 in 1-, 3- and 5-years are 0.667, 0.635, 0.633, 0.725, 0.696 and 0.685, respectively. However, the average AUC of GSE20713 cohort was low, only 0.617. We estimated that it was due to the small size of the data set.

Risk analysis showed the relationship between subgroups, and plotted the distribution results of survival and death states. (Fig. 5: A2-D2, A3-D3). Fig. 5 (A2-D2) showed the distribution of risk score based patients about TCGA and three GSE cohorts. Moreover, the survival status for each patient were demonstrated in the Fig. 5 (A2-D3). The dotted line in the figure divides the 2 groups of patients. It can be seen that the mortality of patients in low-risk areas was generally lower than that in high-risk areas. Especially in TCGA cohort, since there were many cases in this data set, the results should be more reliable. The figure showed that the mortality density in the low-risk group to the left of the dotted line was lower than that in the high-risk group.

Furthermore, the survival curve showed the survival of the 2 groups in more detail. (Fig. 5: A4-D4) In each figure, the two curves plotted the survival of different groups of patients. The figure demonstrated that with the passage of time, the survival number of high-risk patients was significantly lower than that of low-risk patients at the same time.

Besides, the PCA (Fig.5: A5-D5) and t-SNE (Fig.5: A6-D6) plots for BC showed the distribution of different patient groups, and the patients in the 2 groups can be divided clearly.

Then, the univariate and multivariate Cox regression analyses for the risk score and some other clinical features were performed. Whether univariate (Fig. 5: E1-F1) or multivariate aspects (Fig. 5: E2-F2), the risk score has a certain reference value, and can be identified as an independent predictor of survival in BC patients.

Finally, through the visual representation of heat map (Fig. 5 G), the expression differences of three Subtype Prognostic Genes among different subgroups were verified. The results showed that different genes were different among different groups.

### GO, KEGG, and ssGSEA Analysis based on the Risk Model

Based on the risk label, the Wilcoxon Test method can be used to analyze the difference between the high- and low-risk subgroups. The results showed that 94 genes called **Subgroup-DEGs** in TCGA were different, and they were applied to GO and KEGG enrichment analysis. GO analysis (Fig.6: A-B) showed that the main Biological Processes (BP) of these differentially expressed genes were epithelial tube morphogenesis and urogenital system development, and the main Cellular Component (CC) was collagen-containing extracellular matrix. The main Molecular Function (MF) was the extracellular matrix structural component. KEGG analysis (Fig.6: C-D) showed that these **Subgroup-DEGs** were mainly enriched in the estrogen signaling pathway.

**Figure 6.**
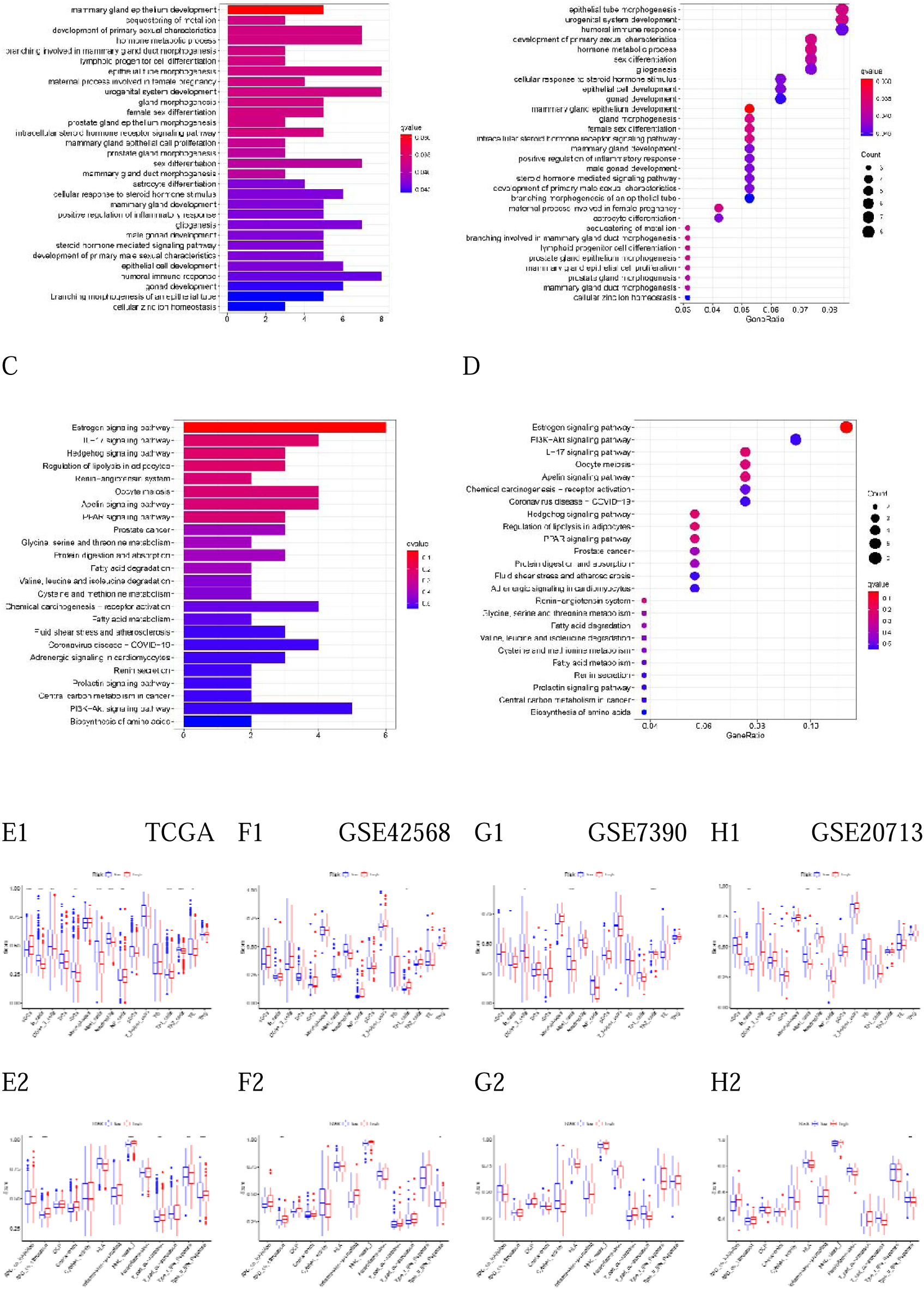
GO, KEGG, GSEA and ssGSEA analysis based on the prognostic model. **A**, **B**, **C**, **D**: Gene Ontology (GO) enrichment analysis and Kyoto Encyclopedia of Genes and Genomes (KEGG) pathway analysis between low- and high-risk group in the TCGA cohort. **E1**, **E2**, **F1**, **F2**, **G1**, **G2**, **H1**, **H2**: Comparison of the enrichment scores of 16 types of immune cells and 13 immune-related pathways between low- and high-risk group in the TCGA, GSE42568, GSE7390, GSE20713 cohorts.

Furthermore, the ssGSEA analysis was applied not only to TCGA cohorts, but also to 3 GEO cohorts. It should be noted that ’***’ indicates p <0.001, ’**’ indicates p <0.01, and ’*’ indicates p <0.05. In the results of *p* <0.05 (Fig.6: E1-H1), the TCGA cohort’s immune scores of low-risk subgroup were higher than that in the high risk subgroup except for the aDCs and Th1 cells. Besides, the low-risk group of GSE7390 and GSE20713 cohorts had higher immune scores, while the GSE42568 cohort’s Th1 cells of high-risk subgroup had a higher immune score.

In the immune pathway analysis with *p* <0.05 (Fig.6: E2-H2), TCGA cohort showed that except for Type II IFN Response, other immune functions had higher immune scores in the low-risk subgroup.

The GSE42568 cohort showed that APC co stimulation had a higher immune score in the high-risk subgroup, while Type II IFN Response had a higher immune score in the low-risk subgroup. In addition, there was no result of *p* <0.05 in GSE7390, but the immune score of low-risk subgroup was slightly higher overall. In the GSE20713 cohort, only *p* value of Type II IFN Response was less than 0.05, which indicated the low-risk subgroup had a higher immune score. It should be noted that the three GEO cohorts have less reliable immune cells and immune pathways, which is due to less data sets.

## Discussion

The gene expression profile of BC can be used to express the heterogeneity of BC at the genomic level. What is most significant is that it can support the establishment of a prognostic model and play a guiding role in the treatment of early breast cancer patients. Nowadays, BC has caused great distress to women’s health. Therefore, it is of great clinical significance to analyze the prognosis of patients through genetic testing.

Necroptosis, morphologically similar to necrosis, is a programmed, caspase-independent form of cell death, which executed by the receptor-interacting protein kinase 1 (RIP1), RIP3, and mixed lineage kinase domain-like protein (MLKL)^[19–20]^. In addition, necroptosis is considered as a promising new target in cancer treatment, which can promote anti-cancer immune response^[21–23]^. However, the biological role of necroptosis pathway in carcinogenesis and the cancer specific role of programmed cell death still needs to be further studied^[21]^. So far, the mechanism and key molecules of necroptosis in the treatment of breast cancer, as well as the cell death pattern of necroptosis are still being studied^[24]^. In this study, we intend to provide insight into the relationship between the NRGs and BC through four aspects of work.

1) **Difference Analysis**. The differences of gene expression between normal people and patients were analyzed. By the Wilcoxon Test, 62 **Different NRGs** were collected, which were regarded as a collection of genes with differences between normal people and patients. To explore the role of Different NRGs, the PPI, correlation network and heat map were analyzed, respectively.
2) **Sub-type Analysis**. 12 **Prognostic NRGs** were screened, and 2 subtypes of BC was identified by these genes. By applying the univariate COX analysis with the cutoff of *p* < 0.05, ’FASLG ’, ’IPMK’, ’TNFRSF1B’, ’PANX1’, ’BCL2’, ’FLT3’, ’PLK1’, ’BACH2’, ’HSP90AA1’, ’LEF1’, ’BNIP3’, ’CD40’ were gathered, which were considered to be characteristic genes related to subtypes. To further study the potential biological characteristics of NRGs, we conducted the Consensus clustering to verify the clustering results, and the survival curve proved that the 2 subtypes did have different survival results. It is worth mentioning that in other studies on BC subtypes, Shuang Shen^[25]^, JianBin Wu et al.^[26]^ divided BC into three categories according to 31 and 8 different pyroptosis-related genes, respectively.
3) **Prognostic Analysis**. It is imperative to establish a reliable prognostic model for optimal clinical outcomes in BC.

In order to achieve this goal, based on the 6 **Same-EGs** "CDKN2A, BCL2, FLT3, PLK1, GATA3 and MYCN", we further obtained three **Subtype Prognostic Genes** (BCL2, FLT3, PLK1) by Cox regression, and obtained the weights by the Lasso Regression, which are -0.02478, -0.22893 and 0.082755, respectively. Using TCGA data as the training set, the threshold value of different subgroups was obtained as the judgment basis of high- and low-risk. Moreover, we found that these three genes were related to NRGs or BC.

As the founding member of the BCL2 (B-cell lymphoma 2) family of regulator proteins, it was the first apoptosis regulator found in any organism^[27]^. Also, as a hot point of PD-1 immunotherapy, it can also maintain a lasting anti-tumor response, and its reactive apoptosis can be resistant to the failure of immunotherapy^[7][28]^. It has also been shown that BCL2 prevents apoptosis and promotes cell survival by neutralizing BH3 proteins^[29]^. Besides, both apoptosis and necrosis can be suppressed by the antiapoptotic BCL2 proteins, and further inhibit mitochondrial outer membrane permeabilization and mitochondrial permeability transition^[30–31]^. When analyzing Necroptosis-Related LncRNAs about gastric cancer, Zirui Zhao et al.^[7]^ Screened 16 LncRNAs in the established prognostic model, including AC012409.3 and AC069549.1, which are related to BCL2. In addition, the expression of BCL2 in BC patients and its impact on the overall survival of patients are also related^[32–35]^.

FLT3 (fms like tyrosine kinase 3) is also known as the Cluster of differentiation antigen 135. It’s a cytokine receptor which belongs to the receptor tyrosine kinase III^[36]^. Internal tandem duplications of FLT3 (FLT3-ITD) are the most common mutations associated with the acute myelogenous leukemia (AML), and it’s a prognostic indicator of related diseases. Moreover, Hillert LK et al. demonstrated that the 32D-FLT3-ITD cells treated with SMAC mimetic bv6 and CD95L can make these cells sensitive to apoptosis and necroptosis^[36]^.

PLK1 (polo-like kinase 1) is also called serine/threonine-protein kinase 13, consists of 603 amino acids, and the molecular weight is about 66 kDa. Some literatures show that PLK1 is associated with BC and plays a transcriptional regulatory role in the interphase of BC cells^[37][38]^.

**4) Sub-group Analysis**. Enrichment analysis and immunoassay were performed on high- and low-risk patients. After building the model, TCGA and GEO cohorts can be assigned corresponding labels to represent the high- and low-risk of patients. We performed the Wilcoxon Test to find out the different genes that lead to high- and low-risk differences. Based on these differential genes, GO and KEGG analyses were applied to TCGA cohort, and ssGSEA analysis was applied to GEO and TCGA cohorts.

In BP analysis, ’adjust-p’ less than 0.05 were ’vegetativeal system development’ (adjust p = 0.041782393) and ’epithelial tube morphogenesis’ (adjust-p = 0.041782393). The corresponding genes were ’LRP2, AGTR1, WNK4, BCL2, AR, ESR1, FOXA1, GATA3’ and ’LRP2, WNK4, BCL2, AR, ESR1, FOXA1, GATA3, PGR’, respectively. BC has also been documented to be associated with these two biological processes. In terms of symptoms, Lester JL and Bernhard LA believed that BC survivors usually have symptoms similar to atrophic vaginitis ^[39]^, and this GO analysis provides a deeper understanding at the genetic level. Besides, eleven genes in CC analysis were associated with ’collagen-containing extracellular matrix ’, they were ’CILP, SPARCL1, CPA3, ADAMTS15, MFAP4, S100A7, THSD4, S100A8, OGN, S100A9, COL14A1’, the ’adjustment-p’ was 0.001417348, which was considered to be associated with Necrosis^[40]^.

In KEGG analysis, 6 genes were included in the ’Estrogen signaling pathway’ (p = 0.000259, q = 0.039461), their corresponding ID number was ’596, 7031, 2099, 51806, 5241, 3868’, which was considered to be related to breast cancer^[41]^. It is worth mentioning that in the KEGG analysis, four genes were considered to be associated with the ’Coronavirus disease-COVID-19’, and their ID numbers were ’ 3572, 185, 730, 4312 ’, but the results were less reliable (p = 0.064236, q = 0.529567603).

In immunocyte analysis, the ADCs (high = 0.493469, low = 0.462574) and Th1 (high = 0.273432, low = 0.247601) contents in the high-risk group were higher, but there was still a large risk. Pondé N et al. also proposed that in the existing research at this stage, ADC has certain limitations in the treatment of BC^[42]^. Zhao X et al. believed that the balance between Th1 and Th2 is an important factor to obtain better immune effect^[43]^. Moreover, excessive Th1 in high-risk groups may have a negative immune effect and thus increase the risk.

In immune pathway analysis, only the low-risk group of Type II IFN Response had higher scores (low = 0.571267753, high = 0.525963672). Cekay et al. proposed that under certain conditions, the high expression of Type II IFN Response could induce cancer cell necrosis^[44]^. Moreover, high Type II IFN Response scores in low risk groups may reduce the risk of BC patients.

There are still several concerns needed to address in our study. First, although we have classified the BC into 2 subtypes, the meaning of each subtype and the corresponding biological differences between subgroups are still not yet fully elucidated, and more detailed verification is required. Secondly, the verification performance of individual GEO is not good enough, and we estimate that the problem comes from the sample size. As for the sample size, TCGA and geo data are not sufficient, and more data are still needed to verify the prognostic model. Thirdly, regarding three NRGs we collected, further clinical verification of their impact and degree on BC is still needed.

## Summary

In all, our study screened 62 different NRGs between normal subjects and BC patients, divided two subtypes of BC according to 12 Prognostic NRGs. More importantly, we screened 3 Subtype Prognostic Genes from NRGs, namely BCL2, FLT3, and PLK1, which were regarded as an independent risk factor of BC prognoses. Based on above, we constructed a prognostic model, which may be able to guide the classification of patients’ prognosis. As a supplement, the different genes in 2 subgroups were studied. The results demonstrated that patients with high-risk BC based on the model showed worse clinical outcomes, and vice versa.

## Data Availability

All data produced in the present study are available upon reasonable request to the authors

## Supplementary

**Table S1.** 67 Necroptosis-Related Genes.

|  |  |  |  |  |
| --- | --- | --- | --- | --- |
| FADD | IPMK | SQSTM1 | HAT1 | RNF31 |
| FAS | ITPK1 | STAT3 | SIRT2 | IDH1 |
| FASLG | SIRT3 | DIABLO | SIRT1 | IDH2 |
| MLKL | MYC | DNMT1 | PLK1 | KLF9 |
| RIPK1 | TNFRSF1A | CFLAR | MPG | HDAC9 |
| RIPK3 | TNFSF10 | BRAF | BACH2 | HSP90AA1 |
| TLR3 | TNFRSF1B | AXL | GATA3 | LEF1 |
| TNF | TRAF2 | ID1 | MYCN | BNIP3 |
| TSC1 | PANX1 | CDKN2A | ALK | CD40 |
| TRIM11 | OTULIN | HSPA4 | ATRX | BCL2L11 |
| CASP8 | CYLD | BCL2 | TERT | EGFR |
| ZBP1 | USP22 | STUB1 | SLC39A7 | DDX58 |
| MAPK8 | MAP3K7 | FLT3 | SPATA2 | TARDBP |
| APP | TNFRSF21 |  |  |  |

**Table S2.** Abbreviations of proper nouns.

| Abbreviation | Full name |
| --- | --- |
| NRGs | Necroptosis-Related Genes |
| Different NRGs | Different Necroptosis-Related Genes |
| Subtype-DEGs | Subtype Different Expressed Genes |
| Subgroup-DEGs | Subgroup Different Expressed Genes |
| Prognostic NRGs | Prognostic Necroptosis-Related Genes |
| NR BCGs | Necroptosis-Related Breast Cancer Genes |
| Same-EGs | Same Expressed Genes |

