## Supplemental Tables for "Identification and Modeling of Necroptosis-Related Genes Associated with the Prognosis of Breast Cancer": instruction.txt

1. "cluster.xls" is the cluster result of the two subtypes in Breast Cancer.
2. "clusterDiffGeneExp.xls" is the differential gene expression matrix of these two subtypes.
3. "tcga.uniCox.xls" is the 3 Subtype Prognostic Genes.
4. "GSE20713Risk.xls" is the risk score of GEO cohort in GSE20713.
5. "GSE42568Risk.xls" is the risk score of GEO cohort in GSE42568.
6. "GSE7390Risk.xls" is the risk score of GEO cohort in GSE7390.
7. "TCGARisk.xls" is the risk score of TCGA cohort.
